# Early prediction of renal graft function: Analysis of a multi-centre, multi-level data set

**DOI:** 10.1101/2021.01.04.20248473

**Authors:** Arturo Blazquez-Navarro, Chris Bauer, Nicole Wittenbrink, Kerstin Wolk, Robert Sabat, Chantip Dang-Heine, Sindy Neumann, Toralf Roch, Patrizia Wehler, Rodrigo Blazquez-Navarro, Sven Olek, Oliver Thomusch, Harald Seitz, Petra Reinke, Christian Hugo, Birgit Sawitzki, Nina Babel, Michal Or-Guil

## Abstract

**Introduction:** Long-term graft survival rates after renal transplantation are still moderate. We aimed to build an early predictor of an established long-term outcomes marker, the glomerular filtration rate (eGFR) one year post-transplant (eGFR-1y).

**Materials and Methods:** A large cohort of 376 patients was characterized for a multi-level bio-marker panel including gene expression, cytokines, metabolomics and antibody reactivity profiles. Almost one thousand samples from the pre-transplant and early post-transplant period were analysed. Machine learning-based predictors were built employing stacked generalization.

**Results:** Pre-transplant data led to a prediction achieving a Pearson’s correlation coefficient of r=0.39 between measured and predicted eGFR-1y. Two weeks post-transplant, the correlation was improved to r=0.63, and at the third month, to r=0.76. eGFR values were remarkably stable throughout the first year post-transplant and were the best estimators of eGFR-1y already two weeks post-transplant. Several markers were associated with eGFR: The cytokine stem cell factor demonstrated a strong negative correlation; and a subset of 19 NMR bins of the urine metabolome data was shown to have potential applications in non-invasive eGFR monitoring. Importantly, we identified the expression of the genes TMEM176B and HMMR as potential prognostic markers for changes in the eGFR.

**Discussion:** Our multi-centre, multi-level data set represents a milestone in the efforts to predict transplant outcome. While an acceptable predictive capacity was achieved, we are still very far from predicting changes in the eGFR precisely. Further studies employing further marker panels are needed in order to establish predictors of eGFR-1y for clinical application; herein, gene expression markers seem to hold the most promise.

## INTRODUCTION

Long-term outcomes of renal transplantation are still disappointing, with a median graft survival time of around ten years.[1,2] Currently, a large effort is being undertaken to determine new risk factors for the outcome of renal transplantation in order to improve therapy and organ allocation.[3–6] While a number of risk factors have been identified (including HLA mismatch, age of donor or cold ischemia time), they are still not sufficient for a precise prediction of transplant outcome.[3,4,6–8] Therefore, machine learning-based risk assessment models are envisaged to assist medical decision-making in patient therapy and eventually improve patient care.[9]

The most commonly used end-point for risk assessment models is the incidence of acute rejection.[10–14] However, acute rejection occurs rather infrequently in renal transplantation and treatments against acute rejection are reasonably effective.[1,15–17] The estimated Glomerular Filtration Rate (eGFR) could therefore offer a more appropriate end-point.[6] In fact, the eGFR one year post-transplant (eGFR-1y) is an accepted marker of long-term transplant outcome.[18–20] eGFR is an estimate of renal function calculated based on the concentration of serum creatinine and demographic characteristics of the patient; it results from the combined influence of all individual risk factors.[8,21] This is the reason why complex predictive markers are needed – we cannot expect that only one marker will predict eGFR-1y. However, to our knowledge there exists still no predictive model of eGFR with a sufficient predictive performance for the clinic.

The goal of this work is to develop a predictive model for the eGFR-1y employing data collected in the pre-transplant and early post-transplant period. Furthermore, we aimed to identify and characterize potential novel markers of the renal function. For this, we have acquired a multi-level biomarker panel with an eminent immunological focus. Importantly, the choice of markers was not based on an explorative multi-omics approach but rather on data known for their mechanistic connection to transplant outcome. The markers used here were already identified as useful in previous studies addressing immunological risk factors of acute transplant rejection.[22–28]

The analysis was performed according to the following strategy: First, predictive models of eGFR-1y were calculated for each marker subset and visit. Second, predictive models were combined to provide a final prediction of eGFR-1y. Third, the individual markers of the best predictors were investigated in detail, especially to find out whether these markers were diagnostic (reflecting the renal function at the time of sampling), or prognostic (predicting future changes of the renal function). Finally, a second predictive model was built, using exclusively prognostic markers.

## MATERIALS AND METHODS

### Patient population and monitoring

We characterized the patient cohort of the randomized, multi-centre Harmony trial (NCT 00724022) for a biomarker panel as part of the e:KID study.[17] Inclusion criteria were the availability of follow-up data for eGFR-1y, and for at least one of the three study visits. The study was carried out in compliance with the Declaration of Helsinki and Good Clinical Practice. The trial was approved by the Ethics Committee of the Gustav Carus Technical University Dresden.

As published before, the patients of the patient cohort were treated with a quadruple (arm A) or triple (arms B and C) immunosuppressive therapy.[17] Shortly, arm A received a basiliximab induction therapy and maintenance therapy consisting of tacrolimus (Advagraf®, Astellas), mycophenolate mofetil (MMF) and corticosteroids. Arm B received the same induction and maintenance therapy, with corticosteroids withdrawn at day 8 post-transplant. Finally, induction therapy for arm C consisted of rabbit anti-thymocyte globulin (ATG), with the same maintenance therapy as arm B. Furthermore, based on their risk constellation patients received an anti-cytomegalovirus prophylaxis consisting of valganciclovir.[29]

Patients were monitored for eGFR during the first post-transplant year, calculated using the CKD-EPI formula and measured in mL·min^-1^·1.73 m^-2^.[21] eGFR was assessed at regular intervals: 2nd week, 1st month, 2nd month, 3rd month, 6th month, 9th month, and 12th month post-transplant.

### Characterization of the patient cohort by a biomarker panel

The patients were characterized by a biomarker panel consisting of five marker subsets (Table 1) at three visits: pre-transplant (pre-Tx), two weeks post-transplant (2w) and three months post-transplant (3m). For a detailed description of the methods employed in the characterization, see the Supplementary Methods.

**Table 1.** Description of the five subsets of the biomarker panel of the e:KID study. The number and list of the markers corresponds to those with a data availability such that they could be employed for prediction for at least one visit (for more details see Statistical analysis: Prediction of eGFR). For the definition of the gene expression subset abbreviations, see the Supplementary Methods.

| **Sample type** | **Marker type** | **Markers** | **No. of markers** | **Name of subset** |
| --- | --- | --- | --- | --- |
| Serum | Cytokine concentration | Interleukin 19 (IL-19), Interleukin 22 (IL-22), stem cell factor (SCF), soluble suppression of tumorigenicity 2 (sST2), Beta-defensin 2 (BD2), angiogenin and endostatin | 7 | Cytokines |
| Whole blood | Gene expression of tolerance and rejection markers | CD79B, CD200, CD247, CD274, CXCL10, FCRL1, FCRL2, FOXP3, HMMR, HS3ST1, LAG3, MAN1A1, MS4A1, NAV3, PNOC, SH2D1B, SLC8A1, TCL1A, TLR5 and TMEM176B | 20 | Gene expression |
| Urine | Metabolomic parameters (given as intensities) | Spectral regions | 377 | Metabolome |
| Serum | Single antigen reactivity profile | A subset of HLA-1 allotypes | 97 | SAB |
| Various | Demographic and clinical characteristics | Demographics of recipient (age, weight, sex, BMI and underlying renal disease), donor characteristics (donor age, last measured creatinine, donor type and expanded criteria donor), panel reactive antibodies, previous transplants, cold ischemia time, anti-HLA mismatch (A, B and DR), cytomegalovirus and Epstein-Barr virus mismatch, physical examination values (blood pressure, pulse and temperature). | 56 | Clinical |
|  | Therapeutic strategy | Therapy arm, valganciclovir prophylactic therapy, tacrolimus concentration/dose ratio at 2w, MMF dose at 2w |  |  |
|  | Lab values | eGFR, blood viral load (BK virus, cytomegalovirus and Epstein-Barr virus), anti-BK virus IgG antibodies, blood values (white blood cells, red blood cells, platelets, haematocrit, haemoglobin, alanine transaminase, aspartate transaminase, alkaline phosphatase, c reactive protein, glucose, calcium, phosphate, triglycerides, cholesterol, total protein and albumin). |  |  |

Shortly, the cytokine concentration levels were measured by ELISA. Gene expression markers were selected based on their role in operational tolerance or rejection and measured employing TaqMan Gene Expression Assays (Thermo Fisher Scientific).[25–28] Serum antibodies (SAB) were screened employing HLA-1 mixed antigen bead assay; the raw mean fluorescence intensity for each bead was employed for prediction.[10] The urinary metabolomics spectrum was determined employing nuclear magnetic resonance (NMR), which was binned and normalized to facilitate the analysis. Finally, while viral loads were monitored by qPCR as described before, other clinical data were measured de-centrally and provided by the transplantation centres.[30]

### Statistical analysis: Descriptive statistics

Categorical variables are summarised here as numbers and frequencies; quantitative variables are reported as median and interquartile range (IQR). Box plots depict the median, first and third quartile of a variable; the maximum length of the whiskers corresponds to 1.5 times the IQR. Correlations of two continuous variables were calculated employing Pearson’s correlation coefficient r. For the visual representation of linear relationships between variables, the regression line and the confidence interval were calculated employing Deming regression; the 95% confidence interval is shown as a shaded grey area.

All statistical and modelling analyses were performed employing R (Version 3.6.1).[31]

### Statistical analysis: Prediction of eGFR

Prediction of eGFR-1y was performed employing the method of stacked generalization, which aims to combine several weak predictors to produce a precise final predictive model.[32] Reason for this was the differences in data availability patterns between the single subsets, as few patients had available data for all subsets for a given time point. Thus, a predictor was created for each visit and each one of the five marker subsets.

The marker subsets were employed for the prediction as follows: For each visit and marker subset, only markers with <20% missing values were taken into account. This percentage was calculated considering only the patients (partially) characterized for the marker subset at hand, not the entire cohort. Missing values were imputed employing the R package mice (Version 3.8.0) with the method “classification and regression trees” for multiple imputation with 5 multiple imputations and 50 iterations.[33]

Prediction of eGFR-1y was performed employing elastic net regression (R package glmnet, Version 3.0-2) with cross-validation.[34] The elastic net regression was performed with eGFR-1y as the dependent variable and the marker subset (as defined above) as independent variables. Importantly, elastic net regression performs variable based on its parameters alpha and lambda, so that only part of the measured markers are employed for prediction. To avoid overfitting, the elastic net regression was nested in a leave-p-out cross validation (p=5); thus, each iteration of the elastic net regression was optimized on a training set of n-5 patients (where n is the number of the patients in the subset) and calculated for 5 patients. The values of the elastic net parameters alpha and lambda were optimized for each training set employing the function cva.glmnet (R package glmnetUtils, Version 1.1.5), with 10 cross-validation folds.[35] The vector of possible values for alpha was defined as the cubic potency of an arithmetic progression between 0 and 1, with a common difference of 0.1. The value of the parameters for each training set was set to the combination of alpha and lambda achieving the minimum cross-validated mean error in the training set (alpha.min and lambda.min). The resulting model for each training set was employed for the prediction of eGFR-1y of the corresponding test set. The performance of the predictor was assessed employing the Pearson’s r of the correlation between measured and predicted eGFR-1y.

The final prediction of eGFR-1y for each visit was estimated considering all predictors that achieved r>0.15. This final prediction was calculated for each individual patient as a weighted mean of all the available predictions; the weighs were defined as the estimated Pearson’s correlation coefficient r for each predictor. Importantly, no imputation was performed in this step, so that for each single patient only available predictors were employed. The quality of the final prediction for each of the visits was likewise assessed employing Pearson’s correlation coefficient r.

### Statistical analysis: Selection of important markers and identification of prognostic markers

The selection of markers important for the eGFR-1y prediction from each marker subset was based on the internal variable selection of elastic net regression. In the case of the metabolome prediction, due to the high number of markers, variable selection was performed with a different optimization of lambda. Thus, the value of lambda leading to the most regularized model within one standard error of the minimum cross-validated mean error (lambda.1se) was employed for variable selection of the metabolome predictor (but not for calculation of eGFR-1y). The importance of a marker was defined as the percentage of cross-validation iterations for which the marker was selected in the regression. A predictor achieving an importance of 100% was considered to be important for the prediction.

To compare the performance of two different predictors of eGFR, the differences in the absolute error of each prediction were tested employing the paired samples Wilcoxon signed rank test.

To determine whether a set of markers was prognostic – i.e. associated with eGFR-1y independently of the eGFR at the time of sampling – a multiple linear regression analysis was employed. Thus, the regression model incorporated eGFR-1y as dependent variable and the variable(s) of interest and the eGFR at the time of sampling (eGFR-2w or eGFR-3m) as independent variables. The P values for the independent variables of the regression model were calculated employing the *t* test. Variables achieving a P value below 0.050 were considered to be prognostic for eGFR-1y independently from eGFR at the time of sampling, therefore providing an added value for the prediction of eGFR-1y.

## RESULTS

### Characteristics of the patient cohort

Out of 540 patients, a total of 376 (69.6%) patients from 14 transplant centres were characterized. A total of 958 samples were measured. The characteristics of the patient cohort are presented in Table 2; eGFR dynamics during the follow-up are depicted in Figure 1. An overview of the samples characterized for each marker subset is provided in Table 3. Importantly, the patient cohort (N=376) had significantly higher eGFR at 2w than the Harmony patients lost to follow-up (N=164) (35.5 [19.9-47.6] vs. 25.6 [12.2-39.8] mL·min^-1^·1.73m^-2^; P<0.001).

**Table 2.**
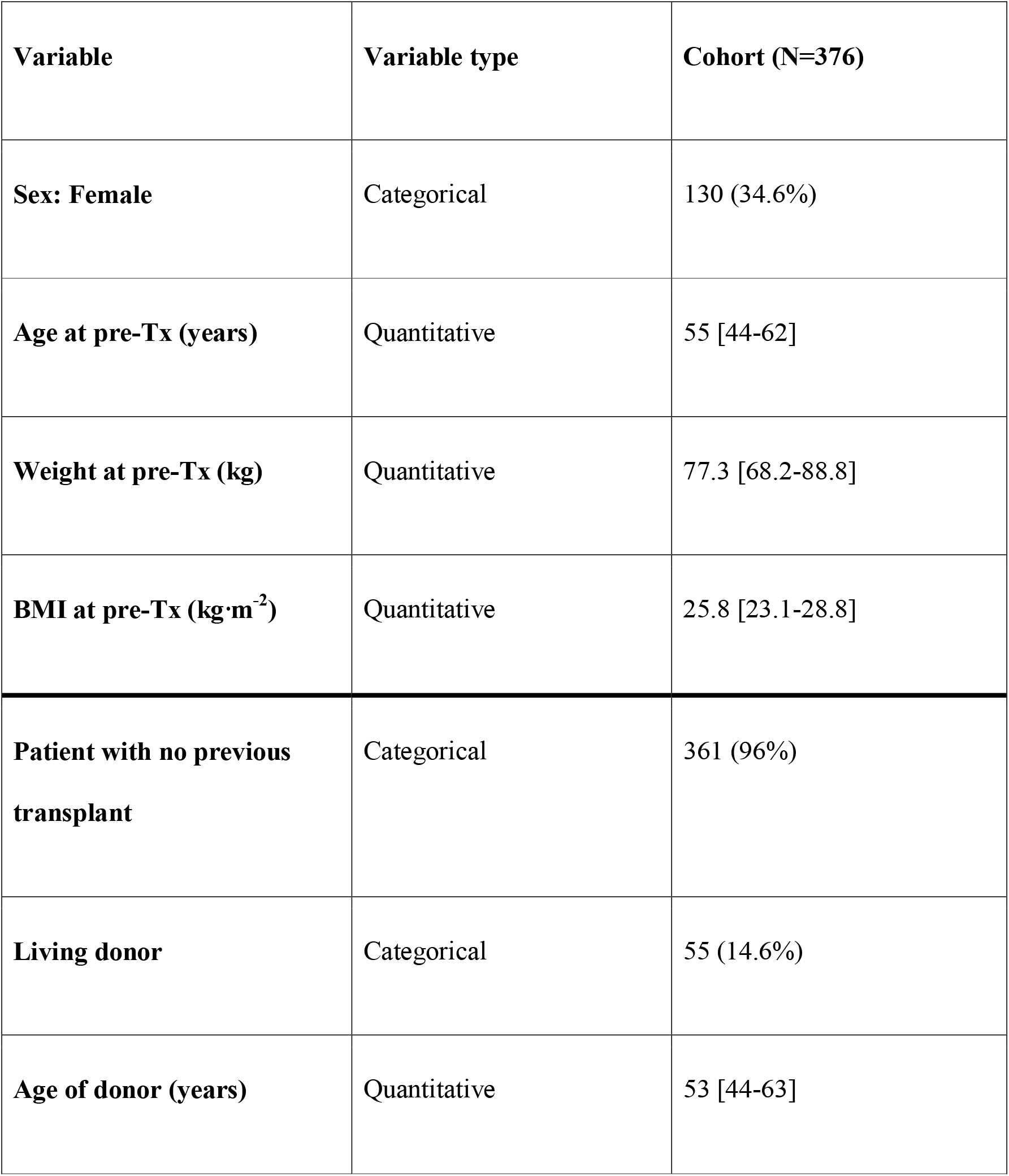
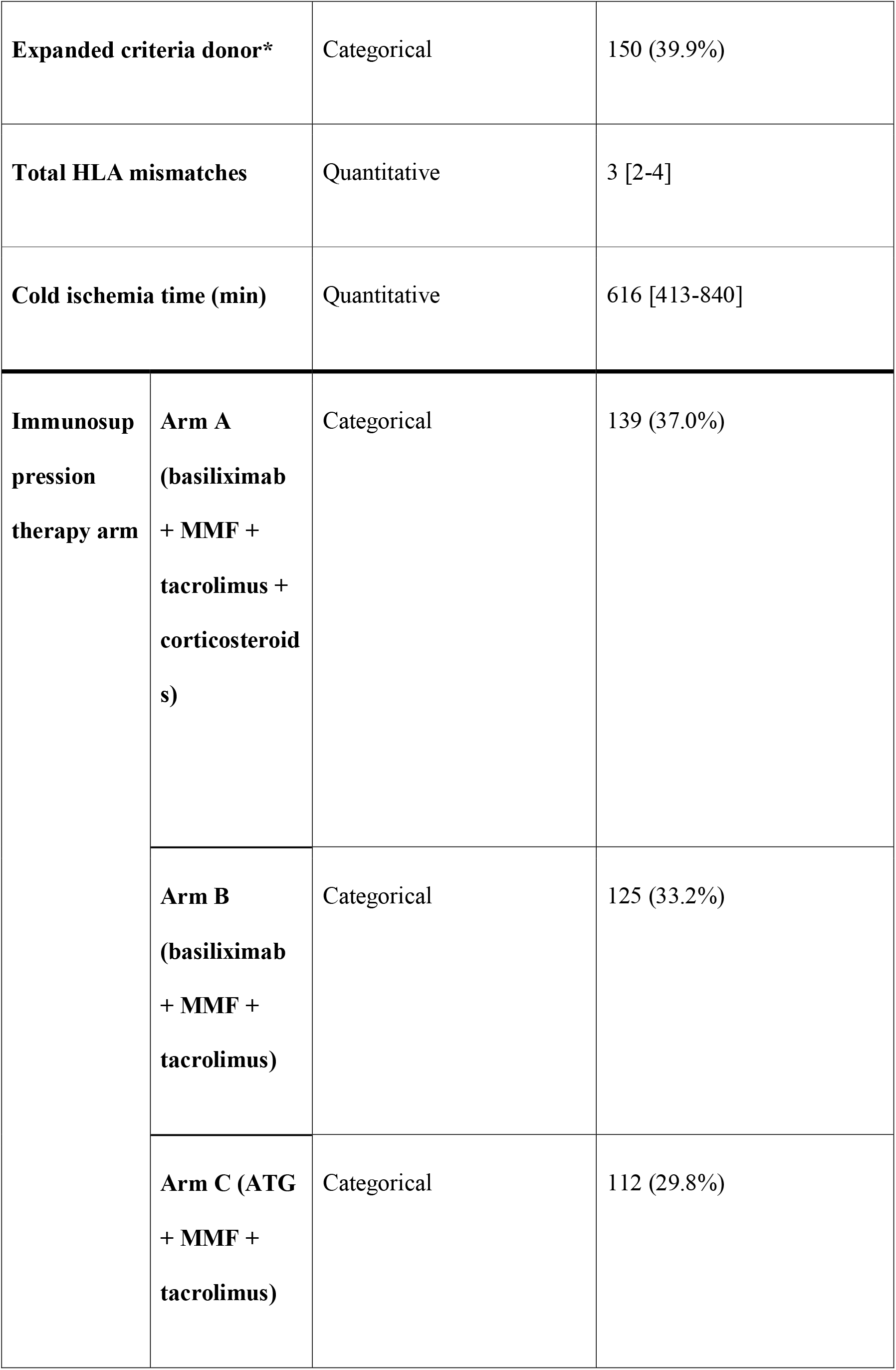
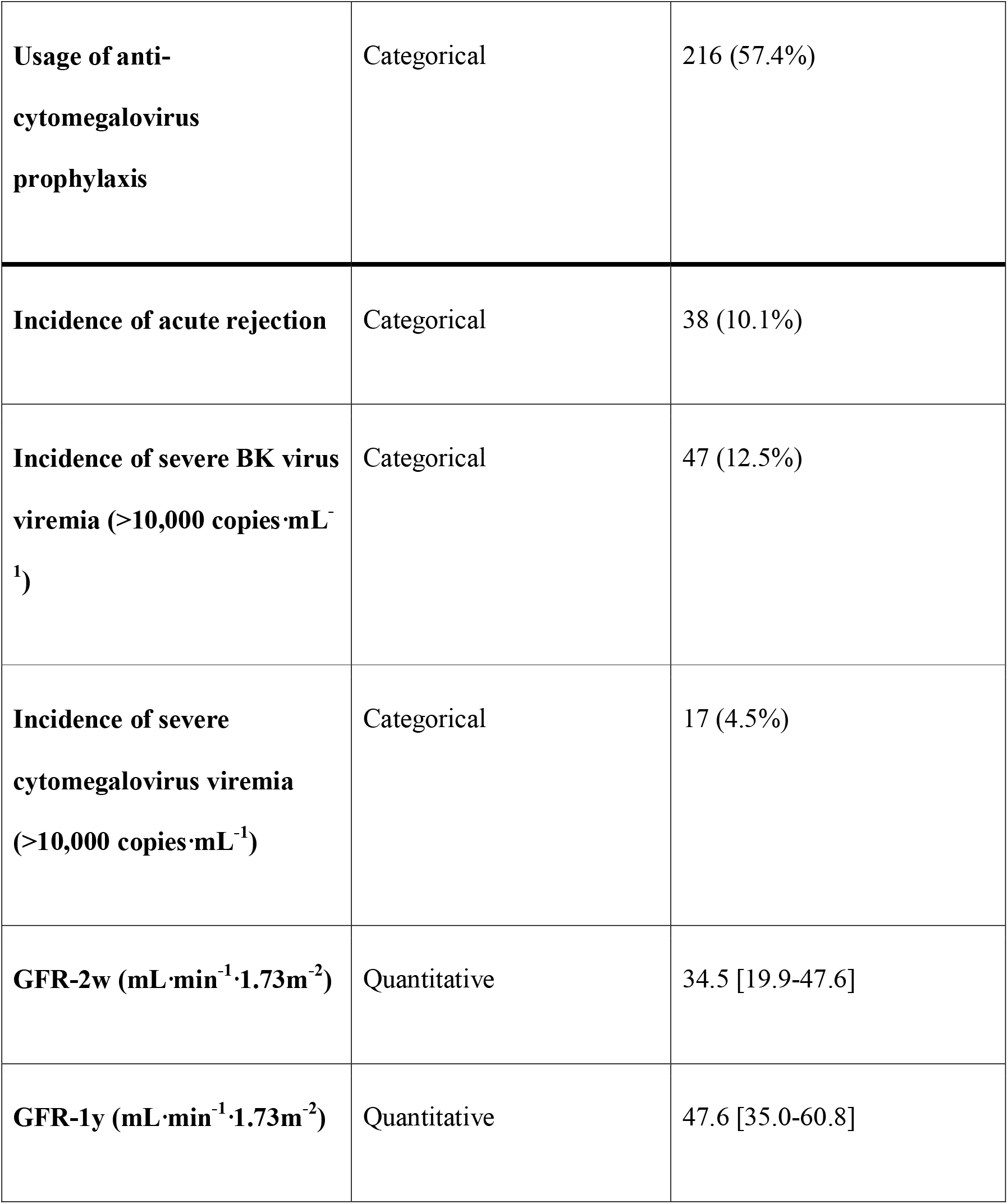
Characteristics of the patient cohort, including demographics, treatment and transplant outcome. Categorical variables are shown as count (% frequency), continuous variables as median [IQR]. *The expanded criteria comprise a donor age greater than 60 years, or an age greater than 50 years combined with at least two of the following factors: cerebrovascular accident as the cause of death, hypertension, or a serum creatinine level of more than 1.5 mg·dL^-1^.[17]

| **Variable** | **Variable type** | **Cohort (N=376)** |
| --- | --- | --- |
| **Sex: Female** | Categorical | 130 (34.6%) |
| **Age at pre-Tx (years)** | Quantitative | 55 [44-62] |
| **Weight at pre-Tx (kg)** | Quantitative | 77.3 [68.2-88.8] |
| **BMI at pre-Tx (kg·m<sup>-2</sup>)** | Quantitative | 25.8 [23.1-28.8] |
| **Patient with no previous transplant** | Categorical | 361 (96%) |
| **Living donor** | Categorical | 55 (14.6%) |
| **Age of donor (years)** | Quantitative | 53 [44-63] |

|  |  |  |  |
| --- | --- | --- | --- |
| **Expanded criteria donor\*** |  | Categorical | 150 (39.9%) |
| **Total HLA mismatches** |  | Quantitative | 3 [2-4] |
| **Cold ischemia time (min)** |  | Quantitative | 616 [413-840] |
| **Immunosuppression therapy arm** | **Arm A**<br>**(basiliximab + MMF + tacrolimus + corticosteroids)** | Categorical | 139 (37.0%) |
|  | **Arm B**<br>**(basiliximab + MMF + tacrolimus)** | Categorical | 125 (33.2%) |
|  | **Arm C (ATG + MMF + tacrolimus)** | Categorical | 112 (29.8%) |

**Table 2.** Characteristics of the patient cohort, including demographics, treatment and transplant outcome.
|  |  |  |
| --- | --- | --- |
| **Usage of anti-cytomegalovirus prophylaxis** | Categorical | 216 (57.4%) |
| **Incidence of acute rejection** | Categorical | 38 (10.1%) |
| **Incidence of severe BK virus viremia (>10,000 copies·mL<sup>-1</sup>)** | Categorical | 47 (12.5%) |
| **Incidence of severe cytomegalovirus viremia (>10,000 copies·mL<sup>-1</sup>)** | Categorical | 17 (4.5%) |
| **GFR-2w (mL·min<sup>-1</sup>·1.73m<sup>-2</sup>)** | Quantitative | 34.5 [19.9-47.6] |
| **GFR-1y (mL·min<sup>-1</sup>·1.73m<sup>-2</sup>)** | Quantitative | 47.6 [35.0-60.8] |

**Table 3.**
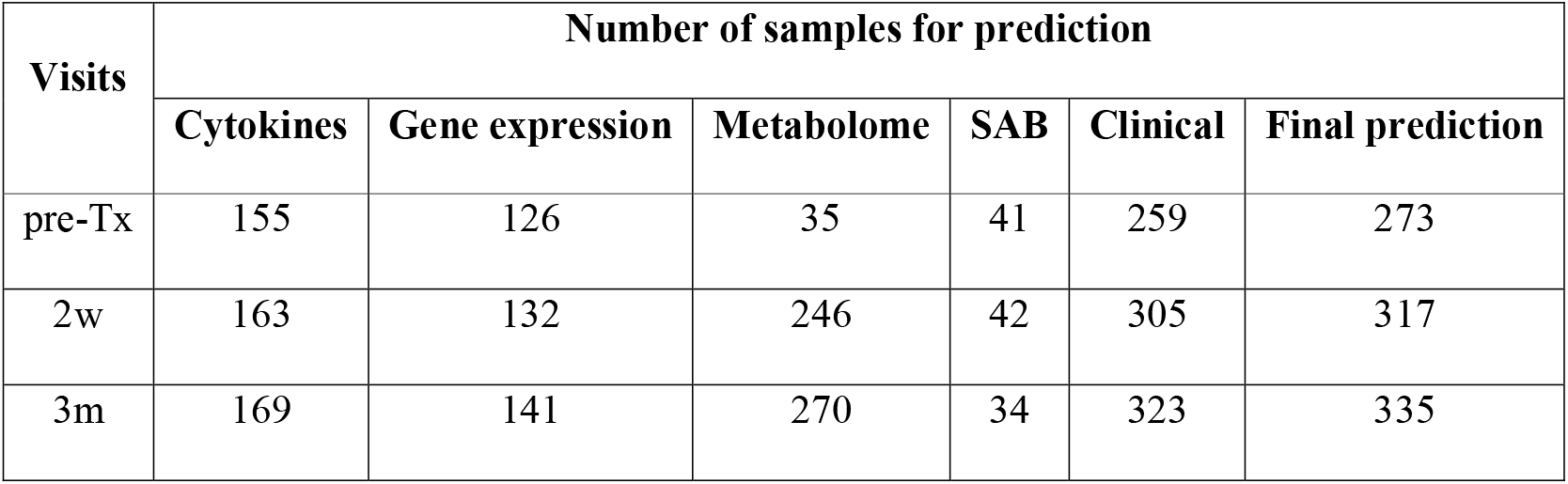
Number of samples employed for the prediction of eGFR-1y for every visit and marker subset, including the final prediction. For the size and composition of the marker subsets, see Table 1.The low number of pre-Tx metabolome samples is due to the low availability of pre-Tx urine samples.

**Figure 1.**
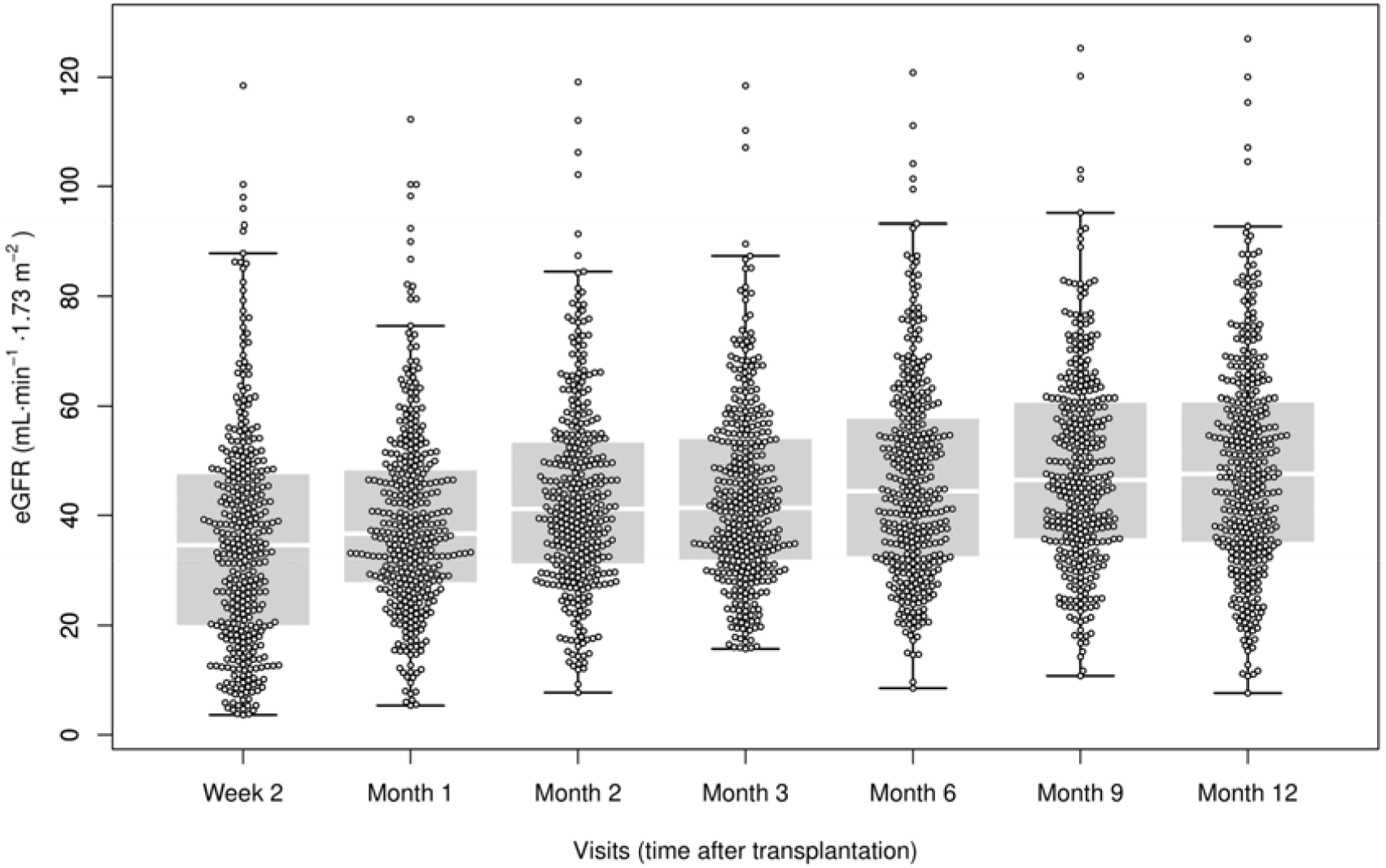
eGFR dynamics in the patient cohort.

### Prediction of eGFR-1y with pre-transplant markers was poor, while post-transplant panels achieved an acceptable performance

Due to different data availability patterns, a predictor was generated for each of the five marker subsets (Table 1); the final prediction was generated through stacked generalization without imputation, using all predictors achieving a Pearson’s correlation coefficient of r > 0.15 between measured and predicted eGFR-1y. The predicted values of the final model at pre-Tx, 2w and 3m are shown in Figure 2A. Figure 2B summarizes the results achieved for each marker subset. A detailed representation of the results for each individual marker subset is shown in Figure S1. Importantly, accuracy of prediction increases with time: While the correlation of measured eGFR-1y and predicted eGFR-1y employing pre-Tx markers was poor (r=0.39), already at 2w r was 0.63 and at 3m, r=0.76. The improvement in the prediction associated with time was important, achieving a significantly lower absolute error of the prediction (pre-Tx vs. 2w: P=0.004; 2w vs. 3m: P<0.001).

**Figure 2.**
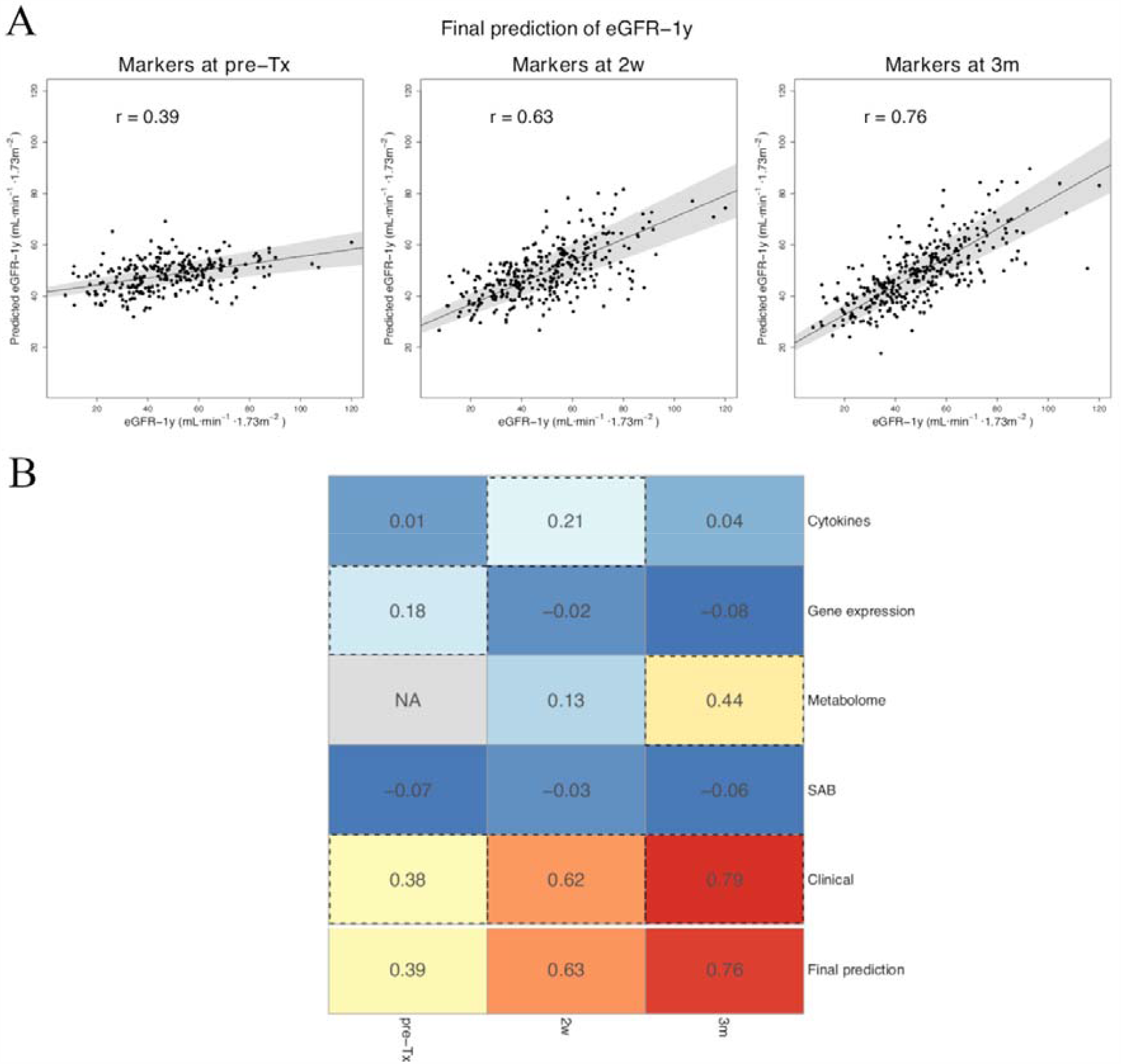
Summary of the prediction of eGFR-1y. (A) Predicted values of the pre-Tx, 2w and 3m stacked models against the experimental eGFR-1y values. (B) Summary of the correlation achieved between the predicted and measure eGFR-1y by each predictor at the three visits. The numeric value indicates the value of the Pearson correlation coefficient r between the predicted and the measured eGFR-1y. Predictors achieving an r>0.15 were employed for the final prediction; these are highlighted with a dashed line. Note that no prediction was performed based on metabolome data pre-Tx due to the low data availability.

For most marker subsets, no accurate prediction could be achieved. While the clinical subset had the highest performance of all predictors, only gene expression at pre-Tx, cytokines at 2w and metabolome at 3m achieved a sufficient quality of prediction to be considered in the final model (see dashed boxes in Figure 2B). To better understand which individual markers are most important for prediction of eGFR-1y, we performed elastic net-based variable selection for these best-performing six predictors. The results are shown in Table S1.

### The pre-transplant immunological state was prognostic of eGFR-1y

We first investigated the two predictors employing pre-transplant (pre-Tx) data: the clinical and the gene expression subsets (see Table S1). For the latter, we identified a complex signature of nine genes as important markers for prediction: CD200, MAN1A1, HS3ST1, NAV3, Foxp3, TCL1A, HMMR, PNOC, and TMEM176B. For the first, we identified eight important clinical markers, comprising: age of donor and recipient, body mass index of recipient, HLA (A, B and DR) donor-recipient-mismatch, cytomegalovirus (CMV) risk constellation and, remarkably, the use of anti-CMV prophylactic strategy.

### Already at the second post-transplant week, eGFR was a highly precise marker for eGFR-1y

We further investigated the prediction of eGFR based on 2w and 3m data. For both visits, eGFR was an important marker for eGFR-1y (Figure 3).

**Figure 3.**
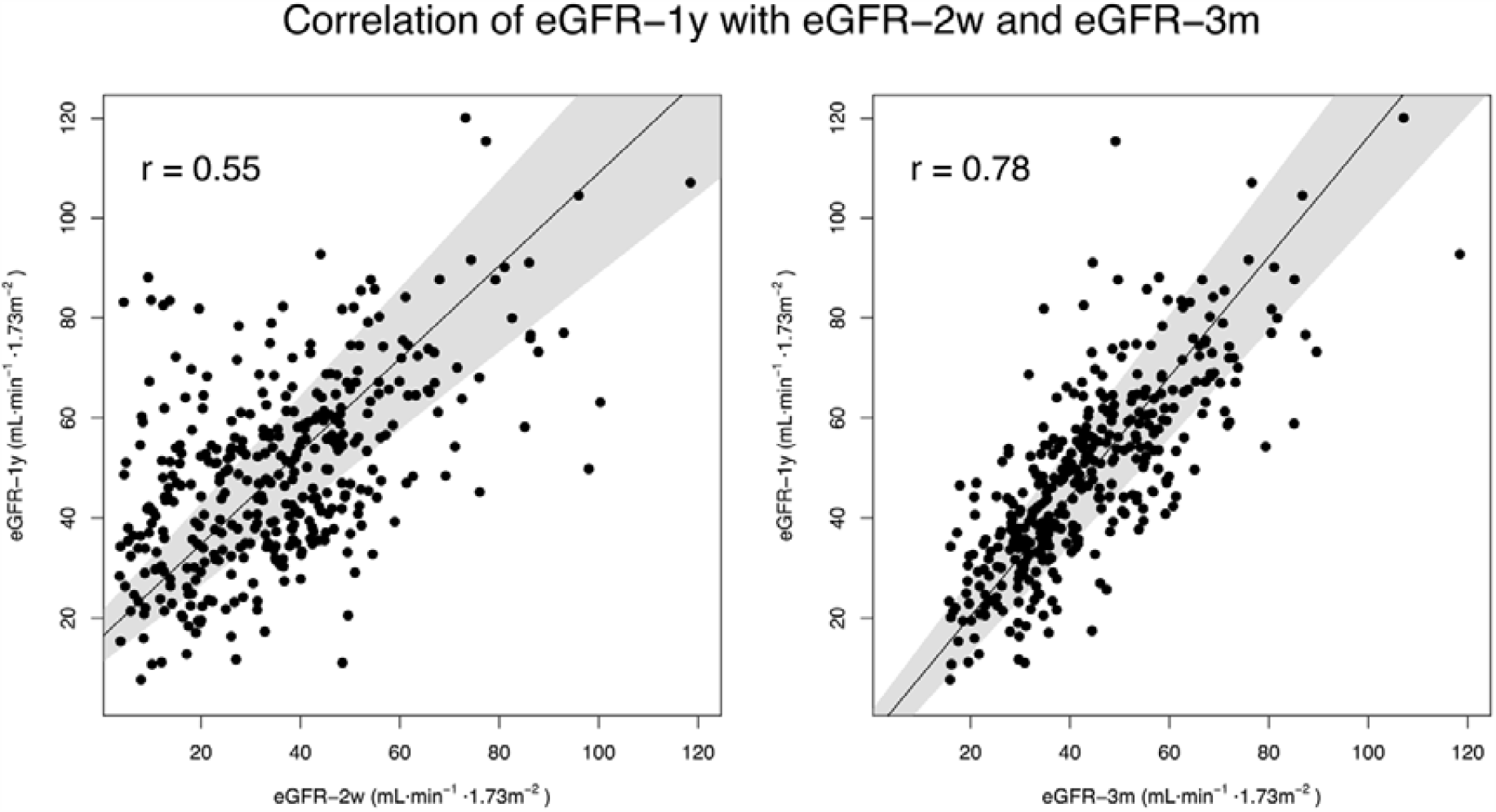
Correlation of eGFR-2w and eGFR-3m with eGFR-1y. The strong correlation of eGFR-2w and eGFR-3m with eGFR-1y demonstrates a high degree of stability of eGFR.

At 2w, we also identified the recipient age, donor age and donor CMV serostatus as important markers, while for 3m, BMI and donor age were identified.

eGFR demonstrated a remarkable stability, achieving alone similar results as the complete clinical marker subset. We thus investigated whether the demographic factors provide an added value for the prediction of eGFR-1y, compared to eGFR alone. Indeed, a multi-parameter regression revealed that recipient age, donor age and donor CMV serostatus at 2w were independently associated with eGFR-1y (recipient age: P=0.045; donor age: P<0.001; donor CMV serostatus: P=0.004; eGFR-2w: P<0.001). Likewise, the association of donor age and BMI at 3m with eGFR-1y was independent from eGFR-3m (donor age: P<0.001; BMI: P=0.008; eGFR-3m: P<0.001). We therefore considered these factors to be prognostic.

### Serum concentration of the stem cell factor (SCF) was negatively correlated with renal function

We examined the cytokine predictor at 2w. Only one cytokine was detected as an important marker by variable selection, the stem cell factor (SCF). SCF at 2w had a negative correlation with eGFR-1y (r=-0.33, P<0.001; Figure 4).

**Figure 4.**
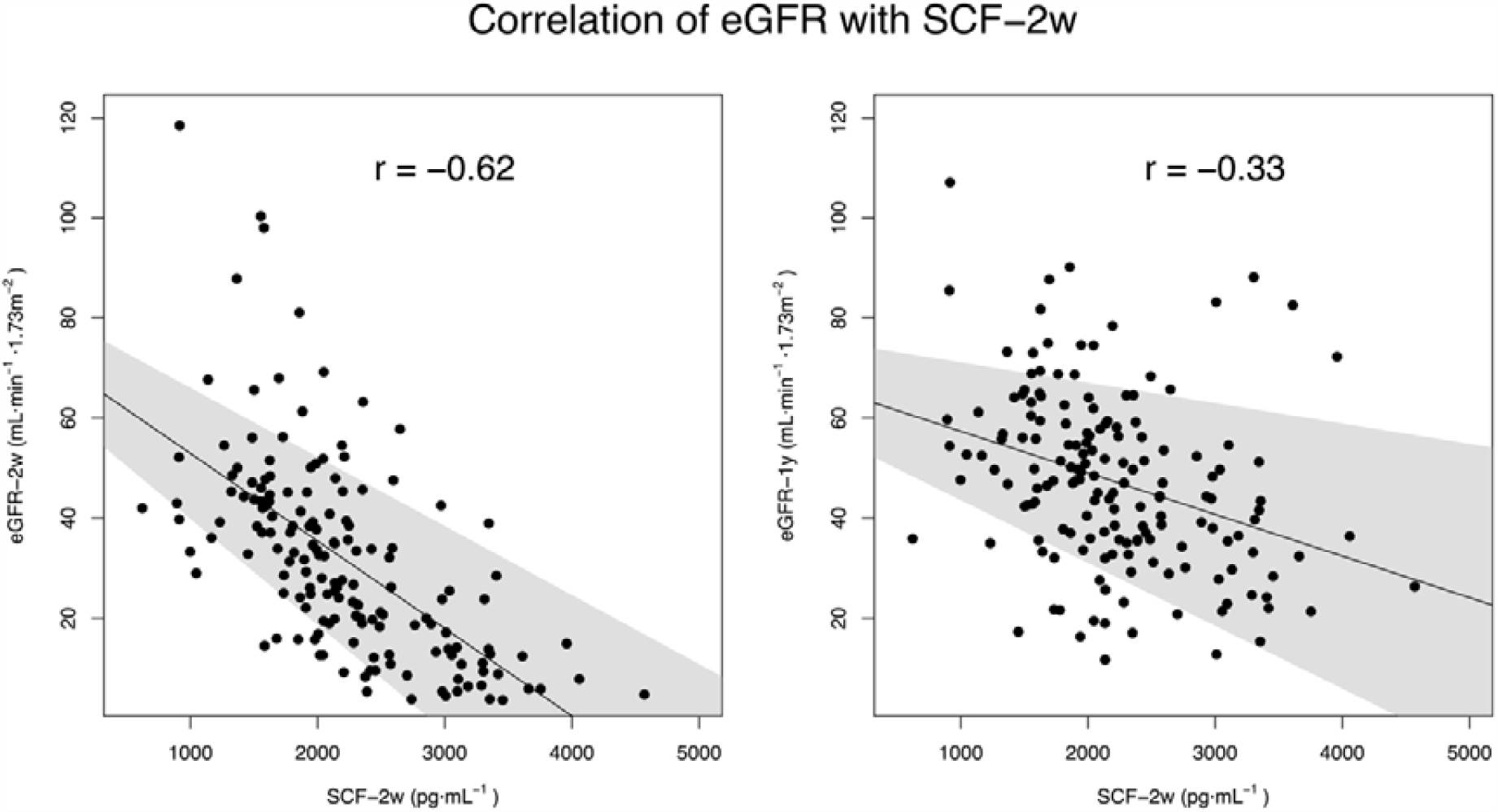
Correlation of eGFR with SCF-2w. Importantly, the correlation with eGFR-2w was stronger than with eGFR-1y.

We further evaluated whether SCF is prognostic for eGFR. As SCF-2w correlated with eGFR-2w stronger than with eGFR-1y (r=-0.62, P<0.001; Figure 4), a prognostic quality of this marker seems unlikely. Furthermore, a bivariate linear regression analysis did not reveal a contribution of the cytokine on the prediction of eGFR-1y (SCF-2w: P=0.612; eGFR-2w: P<0.001). Interestingly, a correlation between SCF and eGFR was observed also at 3m (r=-0.49, P<0.001), suggesting a stable diagnostic accuracy for eGFR.

### eGFR was associated with a urine metabolome signature

Regarding the prediction of eGFR-1y based on the metabolome subset, a complex signature of 19 NMR bins was identified (Table S1). These NMR bins represent up to 35 metabolites, with a median of 2 [1.5-5] candidate metabolites per bin (Table S2). Interestingly, among these candidates, products of amino acid metabolism and creatinine could be found (Table S2).

The presence of creatinine suggests that the urine signature was diagnostic for eGFR-3m, rather than prognostic for eGFR-1y. As expected, a significantly improved prediction (P<0.001) was achieved for eGFR-3m (see Figure 5), compared to the eGFR-1y prediction.

**Figure 5.**
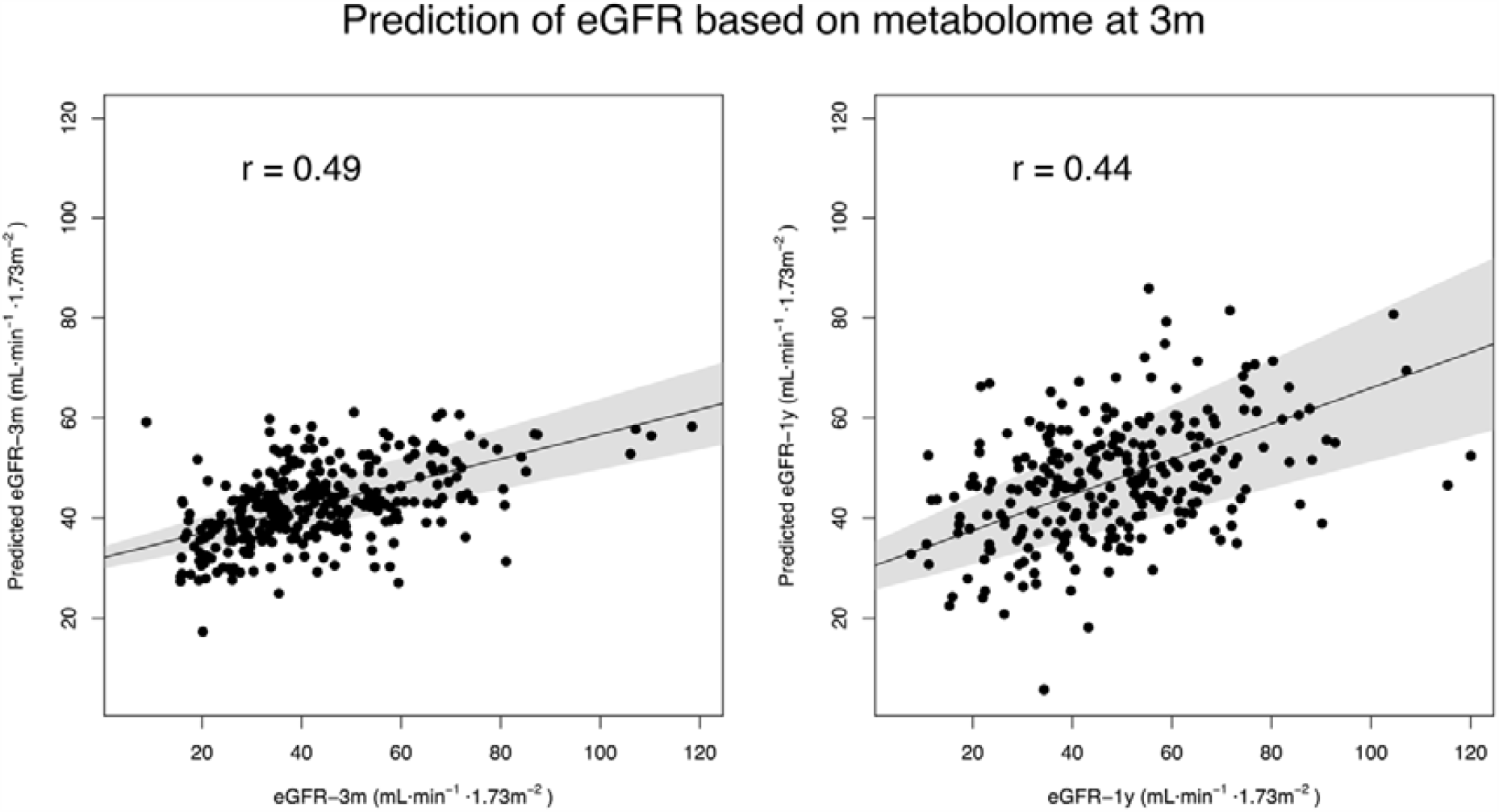
Prediction of eGFR employing the metabolome at 3m. As it can be seen, the quality of prediction achieved for eGFR-3m was higher than for eGFR-1y.

We furthermore analysed whether a predictive advantage for eGFR-1y could be achieved employing together eGFR-3m and metabolome markers at 3m. However, a bivariate linear regression analysis revealed no clear added value of the metabolome predictor, while the effect of eGFR-3m remained highly significant (Metabolome signature: P=0.097; eGFR-3m: P<0.001).

### TMEM176B and HMMR expression at the second post-transplant week was prognostic for changes in eGFR

We further investigated whether any additional prognostic markers can be found in our biomarker panel. With this goal, we repeated the analysis incorporating the eGFR at the sampling point as a variable to the cytokines, gene expression, metabolome and SAB subsets (Figure 6). While no clear improvement of the prediction capacity was achieved in comparison to the main analysis – except perhaps for the gene expression predictor – we identified two potential prognostic markers for eGFR-1y: the expression of the genes HMMR and TMEM176B.

**Figure 6.**
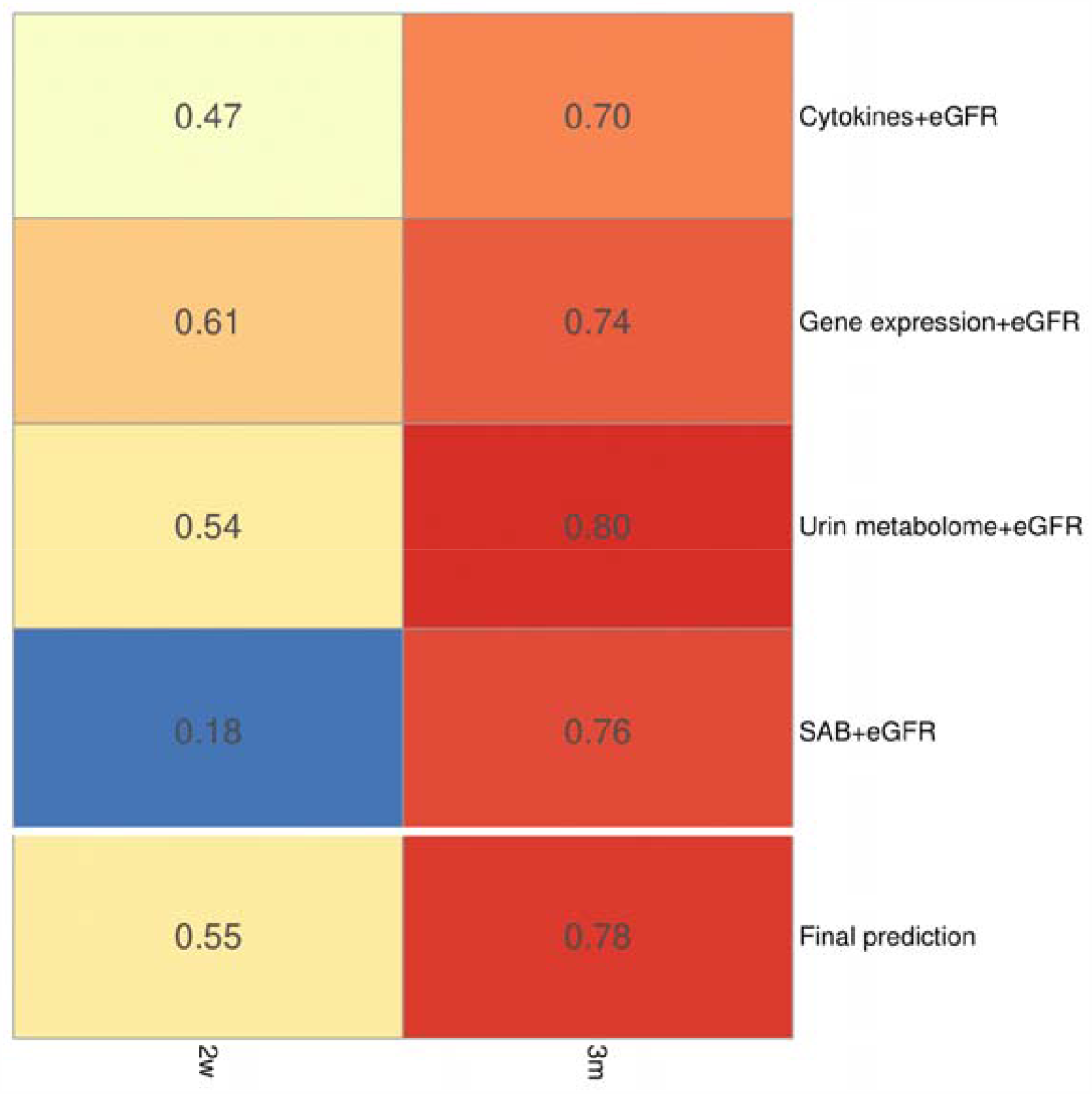
Summary of the analysis for prognostic eGFR-1y models. The figure depicts the correlation achieved between the predicted and measure eGFR-1y by each predictor at the three visits. Each predictor incorporates the eGFR at the time of screening as a factor; all predictors were employed for the final prediction.

High levels of HMMR and TMEM176B were associated with eGFR-1y, independently from eGFR-2w (HMMR: P=0.048; TMEM176B: P<0.001; eGFR-2w: P<0.001; Figure 7). This means that the expression of HMMR and TMEM176B is associated with changes of eGFR between 2w and 1y (see Figure 7). The observed association was not due to a potential correlation with eGFR-2w: None of the two genes showed a significant association with eGFR-2w employing multivariate regression analysis (HMMR: P=0.825; TMEM176B: P=0.057).

**Figure 7.**
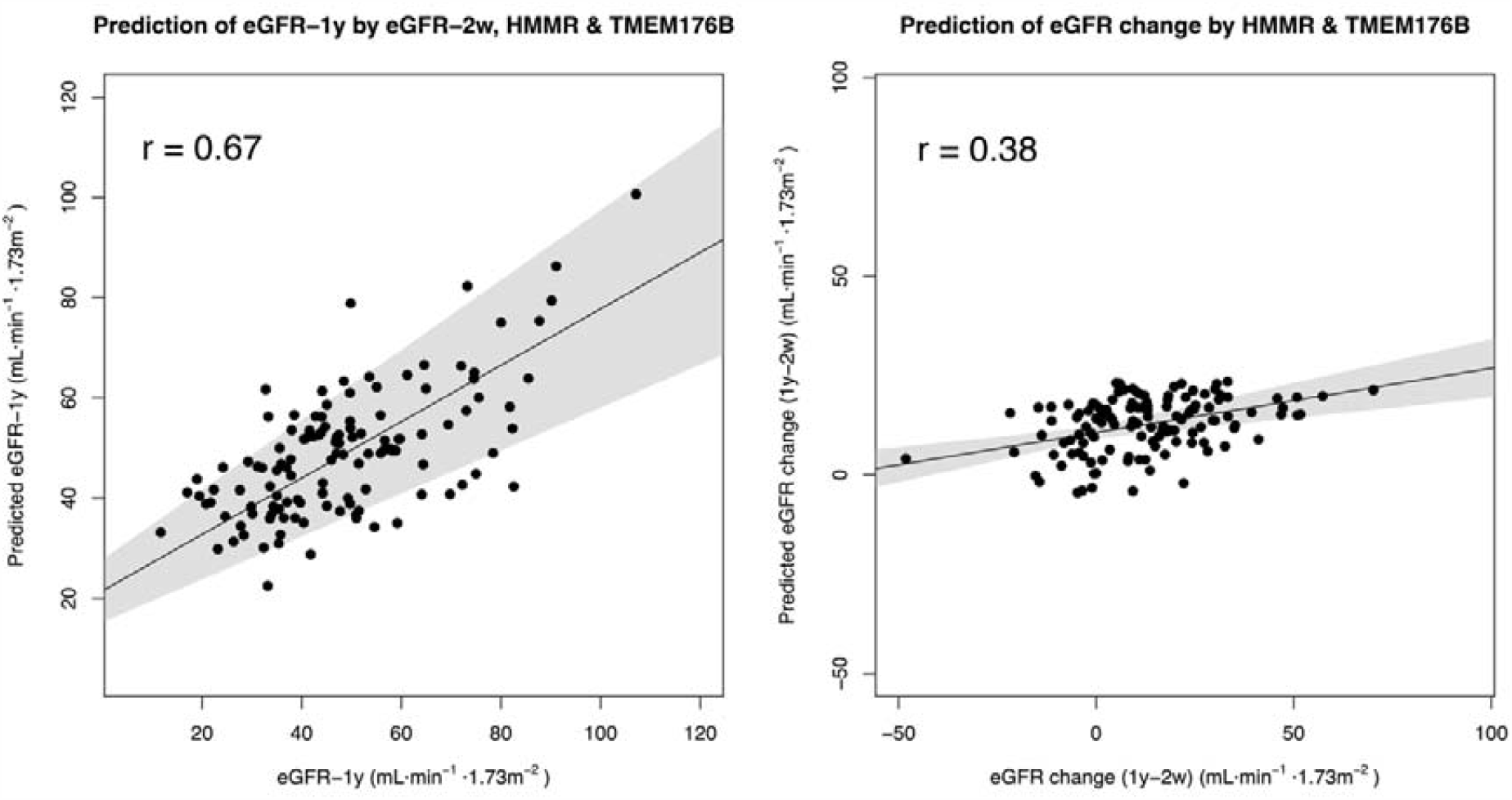
HMMR and TME176B as potential prognostic markers of eGFR. The left panel shows the prediction of eGFR-1y employing eGFR-2w, HMMR and TMEM176B. Note that the achieved correlation is higher than employing eGFR-2w alone (r=0.55, see Figure 5). The right panel shows the prediction of eGFR change between 1y and 2w (eGFR-1y – eGFR-2w) employing only the expression of HMMR and TMEM176B. The prediction shown in this figure was calculated by linear regression without cross-validation, due to the low number of predictors.

## DISCUSSION

In this work, we have performed an in-depth characterization of a large renal transplant cohort in the pre- and early post-transplant period, with the goal of predicting transplant outcome. We have analysed the cohort of the multi-centre Harmony study at three time points using a multi-level biomarker panel, ranging from gene expression to the urine metabolome.[17] Employing these data, we have determined predictive models for the eGFR-1y, identifying potential prognostic and diagnostic markers of the renal function:

- Employing pre-transplant data, the predicted value of eGFR-1y had a correlation of r=0.39 with the measured eGFR-1y. In the post-transplant period, the quality of prediction increased to r=0.63 (two weeks post-transplant) and r=0.76 (three months).
- eGFR demonstrated a remarkable stability in the first post-transplant year: Post-transplant prediction was highly dependent on the value of eGFR at the corresponding time points.
- The gene expression marker subset holds eGFR-1y prognostic value. Especially the genes HMMR and TMEM176B anticipated changes in eGFR after the second post-transplant week.
- Age of donor and recipient, body mass index of recipient, donor-recipient HLA mismatch, CMV mismatch and the use of anti-CMV prophylactic strategy also demonstrated a prognostic character for eGFR-1y.
- The serum concentration of the cytokine SCF had a strong correlation with eGFR at all measured time points, but no prognostic value.
- A complex signature of the urine metabolome with potential applications for the monitoring of eGFR but no prognostic value was identified.

The data of our bio-marker panel represent a milestone in the literature of early assessment of transplant outcomes, as there exists to our knowledge no other study with a comparably profound characterization in a large patient cohort. We have achieved an acceptable predictive capacity employing data from the early post-transplant period. Our results highlight, however, the major challenge of anticipating changes in the renal function: In spite of the large number of markers and samples characterized, we are still far from being able to predict eGFR-1y with an accuracy sufficient for clinical care.

In our results, the eGFR in the early post-transplant period was by far the most precise marker for eGFR-1y. Part of this stability of eGFR can be explained by the nature of our transplant cohort, with low immunological risk and reduced adverse events.[17] This stability is in line with the literature from the last two decades, highlighting that the challenge of post-transplant risk assessment is to anticipate the rare negative changes in eGFR.[36–38]

Here, the most promising markers for eGFR changes belong to the gene expression panel. In the second post-transplant week, the expression of the genes HMMR and TMEM176B anticipated significantly eGFR drops. This observation is coherent with the literature: HMMR is upregulated in lymphocytes during acute rejection in rat.[26] In humans, a study on psoriasis has found a significant association between HMMR and response to immunosuppression.[24] On the other hand, the expression of TMEM176B in peripheral blood is known to be associated with acute rejection in humans.[27] Based on this evidence, we hypothesize that increased expression levels of the genes are a symptom of insufficient immunosuppression, leading to graft inflammation and decrease of eGFR.

Regarding further prognostic markers, the results of our pre-transplant clinical predictor are in basic agreement with the previous work by Lasserre et al.[6] Their work supports the generalizability of our results, as all of the markers identified as important for our predictor were employed in their work for prediction.[6]

We found evidence of an association of eGFR with the cytokine SCF and the urine metabolomic profile. While these associations did not have a prognostic character, i.e. they are markers of eGFR at the time of monitoring, they might still be of interest for achieving a more profound understanding and improved monitoring of eGFR. Thus, there was a strong negative correlation of SCF serum levels with eGFR. An association between SCF and renal function has been observed repeatedly in other contexts.[39–42] Experiments in rat offer a potential cause, suggesting an association of SCF expression with renal fibrosis.[43] Regarding the urine metabolomic profile, our study is a step within current efforts towards a non-invasive monitoring of renal function.[22,44–46] Our preliminary analysis suggests a role of urinary creatinine and amino acid metabolism in the prediction. This is in agreement with the results of Posada-Ayala *et al*. on markers for chronic kidney disease.[46] Importantly, further research is needed for a reduction of the number of candidate metabolites of our signature, to develop a non-invasive renal function monitoring assay.

Our study has some limitations. Firstly, eGFR is only an approximation of the renal function.[21] Ideally, the end-point of a predictive model should be measured directly, employing a gold standard method.[47] Secondly, the composition of the cohort is not representative for the clinical reality, as only patients with low immunological risk were admitted into the study.[17] Thirdly, the study design makes it especially sensitive to survivor bias, as only patients with a (at least partially) functional graft one year post-transplant are evaluated. This is important, since the characterized patients had significantly higher renal function two weeks post-transplant than those lost to follow-up. Finally, not all patients could be characterized for each marker subset, due to issues of sample availability. While the employed modelling approach works well with the different data availabilities for each marker subset, an increased data availability could have allowed for a more accurate modelling of eGFR-1y and identification of potential markers.

In summary, we present the results of an unprecedented characterization of a large renal transplant cohort during the pre-transplant and early post-transplant period, with the goal of predicting the outcome eGFR-1y. Our results highlight the stability of renal function and the difficulty of predicting abrupt changes. They provide two novel candidates for prognostic markers – the expression of the genes HMMR and TMEM176B. Furthermore, we have provided new evidence on the association between renal function and the cytokine SCF and identified a metabolomic signature in urine with potential applications in non-invasive monitoring. Finally, we have achieved an acceptable accuracy in the prediction of transplant outcome, although our capacity to predict abrupt changes in the eGFR still insufficient for a direct application in the clinical context.

## Data Availability

The datasets for this article are not publicly available because the patients have not consented to the publication of their data. Requests for limited access to the data should be directed to Nina Babel.

## FUNDING AND ACKNOWLEDGEMENTS

This work was funded by the German Federal Ministry of Education and Research (BMBF), project e:KID (01ZX1312). The funders had no role in data collection, data analysis, data interpretation, writing of the manuscript, or manuscript submission.

## SUPPLEMENTARY MATERIALS

### Supplementary Methods

**Table S1:** Results of the variable selection for all subsets achieving a prediction with r>0.15. The table shows the name of all markers selected through variable selection, as well as the subset and visit they belong to, their importance in the prediction and the nature of their association with eGFR-1y. The importance is calculated as the percentage of cross-validation iterations for which the marker was considered in the regression. Only markers achieving an importance over 20% are listed here.

**Table S2:** Description of the bins (metabolome at 3m) important for the prediction of eGFR-1y and identification of the candidate molecules for these bins.

**Figure S1:** Detailed results of the prediction of eGFR-1y for every marker subset and visit. The correlation achieved between the predicted and measure eGFR-1y by each predictor is shown. The numeric value indicates the value of the Pearson correlation for the predicted and the measured eGFR-1y.

