## Supplementary Methods for "Early prediction of renal graft function: Analysis of a multi-centre, multi-level data set"

### Monitoring of serum cytokines

Measurement of serum BD2 was performed by an enzyme-linked immunosorbent assay (ELISA) kit from Immundiagnostik (Bensheim, Germany), according to the manufacturer’s instructions. Serum levels of IL-19, IL-22, SCF, angiogenin, endostatin and sST2 were quantified by Quantikine ELISA kits from R&D Systems/Biotechne (Minneapolis, MN, USA), according to the manufacturer’s instructions. The suitability of Quantikine ELISAs for the analyses of patient samples has been confirmed by previous studies.(*1*, *2*)

### Measurement of gene expression markers

We characterized the expression of genes previously identified as upregulated in immunosuppression-free operationally-tolerant kidney transplant such as HS3ST1, SH2D1B, CD79B, MS4A1, PNOC, TCL1A, FCRL1 and FCRL2, or in rejection e.g. HMMR, TLR5, SLC8A1 and VAV3 (for information on the gene abbreviations, see Table A).(*3*–*6*)

Thus, patient blood samples were collected in Tempus Blood RNA Tubes (Thermo Fisher Scientific, Schwerte, Germany) and stored at -20°C until shipment as batches into central immune monitoring lab. RNA was isolated using the MagMAX™ for Stabilized Blood Tubes RNA Isolation Kit (Thermo Fisher Scientific). Up to 1000 ng RNA were transcribed into cDNA using the QuantiTect Reverse Transcription Kit (Qiagen, Hilden, Germany). Gene expression was measured using TaqMan Gene Expression Assays (Thermo Fisher Scientific, see Table 2)*,* microfluidic cards and TaqMan Universal Master Mix (Thermo Fisher Scientific) on the ViiA7 Real Time PCR System (Thermo Fisher Scientific). Reactions were run in duplicates using 384-well microfluidic Custom TaqMan® Array Cards; data were analysed applying the respective ViiA7 Software v 1.2.2. Gene expression of the gene expression marker subset was calculated relative to the median expression of three reference genes (HPRT, B2M and GADPH), using the 2^-ΔΔCt^ method.

| **Gene symbol** | **Gene name** | **Thermo Fisher Assay ID** |
| --- | --- | --- |
| HPRT | Hypoxanthine-guanine phosphoribosyltransferase | Hs02800695_m1 |
| B2M | Beta-2-microglobulin | Hs00984230_m1 |
| GAPDH | Glyceraldehyde 3-phosphate dehydrogenase | Hs99999905_m1 |
| CD79B | CD79B | Hs00236881_m1 |
| CD200 | CD200 | Hs01033303_m1 |
| CD247 | CD247 | Hs00167901_m1 |
| CD274 | CD274 | Hs01125301_m1 |
| CXCL10 | C-X-C motif chemokine ligand 10 | Hs99999049_m1 |
| FCRL1 | Fc receptor like 1 | Hs00957541_m1 |
| FCRL2 | Fc receptor like 2 | Hs00229156_m1 |
| FOXP3 | Forkhead box P3 | Hs00203958_m1 |
| HMMR | Hyaluronan mediated motility receptor | Hs00234864_m1 |
| HS3ST1 | Heparan sulfate-glucosamine 3-sulfotransferase 1 | Hs01099196_m1 |
| LAG3 | Lymphocyte activating 3 | Hs00158563_m1 |
| MAN1A1 | Mannosyl-oligosaccharide 1,2-alpha-mannosidase IA | Hs00195458_m1 |
| MS4A1 | Membrane spanning 4-domains A1 | Hs00544818_m1 |
| NAV3 | Neuron navigator 3 | Hs00372108_m1 |
| PNOC | Prepronociceptin | Hs00918595_m1 |
| SH2D1B | SH2 domain containing 1B | Hs01592483_m1 |
| SLC8A1 | Solute carrier family 8 member A1 | Hs01062258_m1 |
| TCL1A | T cell leukemia/lymphoma 1A | Hs00172040_m1 |
| TLR5 | Toll-like receptor 5 | Hs01019558_m1 |
| TMEM176B | Transmembrane Protein 176B | Hs00962650_m1 |

**Table A.** Detailed information on the reference genes (first three rows) and gene expression subset, including the assay employed for measuring of expression.

### Characterization of urine metabolome

Urine samples were measured by nuclear magnetic resonance (NMR) spectroscopy at numares AG (Regensburg, Germany). Therefore, 600 μl of each urine sample was mixed with *AXINON*® urine additive solution. Insoluble material was removed by centrifugation at 20,000 *g* for 10 min at 20°C. A volume of 600 μl of the supernatant was transferred to 5 mm NMR tubes and kept at 2-6°C until measurement with a Bruker Avance II+ 600MHz NMR-spectrometer and a 5 mm triple resonance TXI probe head (PATXI 1H/D-13C/15N Z-GRD). Before samples were measured in batches of up to 93 samples per run, they were warmed to a target temperature of 37°C in the integrated preheating block. In order to ensure consistent and reproducible measurements, each run included one *AXINON*® urine calibrator sample and two *AXINON*® urine control samples. ^1^H NMR spectra were recorded including pre-saturation to suppress the water signal. Finally, the region containing the residual water signal (4.5 to 5.0 ppm) was excluded from further analysis. The resulting ^1^H NMR spectra were binned in the spectral region of 0.96-9.04 ppm using a bucket size of 0.04 ppm and 50% overlap. Due to the huge concentration differences in urine, these bins were further normalized by dividing them by the total area of the respective spectrum in order to allow the comparison of multiple spectra.

### Screening of HLA-1-specific serum antibodies

Screening of serum antibodies was performed as in Wittenbrink *et al*. 2019.(*7*) Briefly, all sera testing positive for the HLA-1 mixed antigen bead assay and a random subset of negative sera were characterized for HLA-1 specific antigens employing the single antigen bead assay (LABScreen Single Antigen HLA Class I, One Lambda). Both MAB and SAB were performed according to the manufacturer’s instructions: Following incubation at 56°C for 30 min and filtering, 20 µl of undiluted serum were incubated for 30 min with 3 µl of the LABScreen bead mix. After a first washing step (1x LABScreen wash buffer), the mix was incubated with 100 µl of a dilution of PE-conjugated IgG detection antibody for 30 min. After a final washing step, the mean fluorescence intensity corresponding for each bead was measured using a FLEXMAP3D Analyser in combination with xPONENT software version 4.1 (Luminex Corporation, Texas, USA). In this study, the raw mean fluorescence intensity for each bead – without binarization or normalization – was employed to predict eGFR-1y.

### Monitoring and management of viral infection

In several transplant centres, prophylactic approaches using valganciclovir were applied to prevent CMV infection. For our analysis, patients starting a (val)ganciclovir treatment during the first 14 days were considered to have received a prophylactic treatment. Likewise, the provided value for tacrolimus and MMF usage corresponds to visit 2w. In parallel to the clinical monitoring performed at each centre, peripheral blood samples from the eight visits were centrally monitored for cytomegalovirus, Epstein-Barr virus and BK virus by TaqMan quantitative polymerase chain reaction (qPCR), as described previously.(*8*) Briefly, DNA was isolated from serum (BK virus) or whole blood samples (cytomegalovirus and Epstein-Barr virus) according to the manufacturer’s instructions. qPCR was performed employing the Prism 7700 Sequence Detector. The detection level was the lowest viral load measured within the range of linearity.

To determine the amount of anti-BKV IgG antibodies in patient serum, 96 well plates (Costar Assay Platte High Binding, Corning) were coated at 4°C overnight with 1 µg·mL^-1^ of BKV-VP1 protein, which was reconstituted in 100 µl sterile ddH_2_0 to a final stock concentration of 1 mg·mL^-1^. Plates were washed twice with 150 µl wash buffer (Phosphoprotein Detection Wash Buffer, Biorad), blocked with Roti-Block (Carl Roth GmbH) solution for 2 hours at 37°C to minimized unspecific bindings and washed for another two times with wash buffer. Patient serum was diluted 1:250 in Roti-Block solution. The standard curve was prepared using 1:2 serial dilutions of the monoclonal anti-human-BKP-VP1 antibody (Sigma Aldrich) with a starting concentration of 500 ng·mL^-1^. 100 µl of standard and diluted serum samples were then added onto the coated plate and incubated for 1 hour at room temperature. After washing twice, 100µl of 1 µg·mL^-1^ anti-human-IgG-HRP (Abcam) diluted in PBS was added to each well and incubated for 1 hour at room temperature. After washing, the plates were developed using the Substrate Reagent Pack (R&D systems) according to manufactures instructions. Absorbance at 450 nm was measured and anti-BKV-VP1 antibody serum concentrations were calculated by interpolating with the standard curves.
