## Supplementary figures and images for "Early prediction of renal graft function: Analysis of a multi-centre, multi-level data set"

### Figure S1

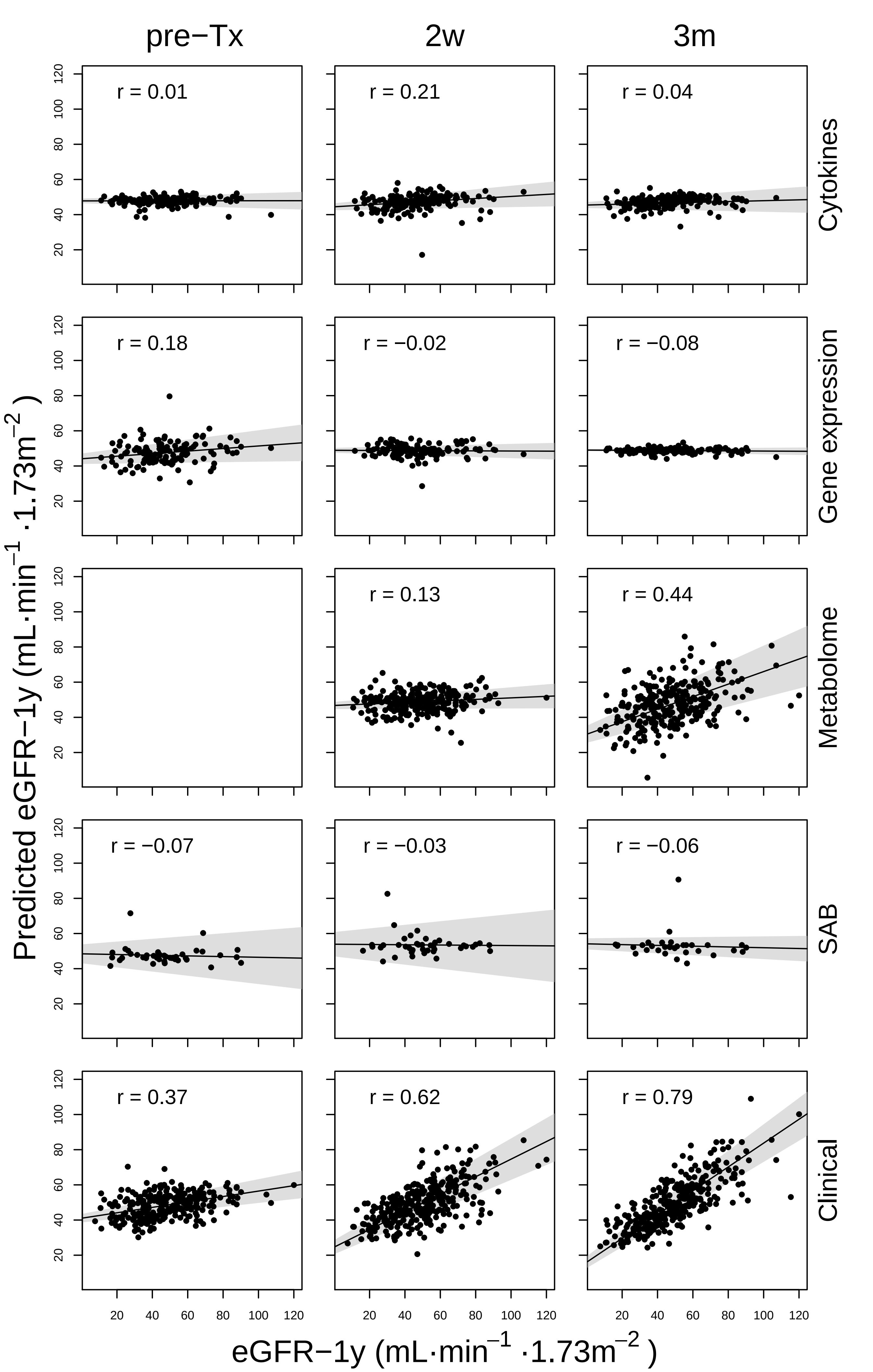
